# Sensitivity and specificity of tuberculosis screening tools in people with diabetes

**DOI:** 10.64898/2026.04.30.26352192

**Authors:** Neha Janrode, Yohhei Hamada, Arshad Taliep, Lauren Barron, Walter Chabaya, Rene T Goliath, Trinh Duong, Amanda Jackson, Shireen Galant, Nashreen Omar-Davies, Louise Lai Sai, Lillian Twentiey, Robert J. Wilkinson, Molebogeng X. Rangaka

**Author notes:** **Corresponding author** Yohhei Hamada, University College London, Institute for Global Health, 3rd floor, Institute of Child Health, 30 Guilford Street, London WC1N 1EH, United Kingdom. Joint first authors (Neha Janrode, Yohhei Hamada).

## Abstract

**Background:** Systematic screening for tuberculosis (TB) is recommended in people with diabetes; however, data on the accuracy of screening tools in this population are lacking. We assessed the accuracy of symptom and chest X-ray screening among people with diabetes.

**Methods:** We consecutively enrolled adults with diabetes attending routine care in South Africa. All participants underwent symptom screening and chest X-ray. A single sputum specimen was collected from all participants and tested by Xpert Ultra. A positive Xpert Ultra result was used as the reference standard.

**Results:** We enrolled 673 participants. The median age was 54 years (interquartile range 47-60 years), and 63.8% were female. HIV prevalence was 17.2%. Prevalent TB was diagnosed in nine participants (1.33%).

Any cough had a sensitivity of 22.2% (95% confidence interval [CI] 2.1-60.0%) and a specificity of 97.5% (95%CI 96.0-98.6%). Expanding the symptom definition to include any of cough, fever, weight loss, or night sweats did not improve sensitivity (22.2 %, 95 % CI 2.1–60.0) and slightly reduced specificity to 96.0 % (95 % CI 94.2–97.0). Chest X-ray abnormalities suggestive of TB demonstrated a sensitivity of 55.6% (95%CI 21.2-86.3%) and a specificity of 95.4% (95%CI 93.4-97.0%). The specificity of chest X-ray was significantly lower in participants with prior TB (87.6%, 95% CI: 79.8–90.6%), compared to 97.2% (95% CI: 95.2–98.5%) in those without (p < 0.01).

**Conclusion:** Symptom-based TB screening has poor sensitivity in people with diabetes. Although chest X-ray improved the sensitivity, it remained suboptimal, with a reduced specificity in people with previous TB.

## Introduction

Tuberculosis (TB) remains a significant global health challenge, with an estimated 10.8 million new cases and 1.25 million deaths in 2023, disproportionately affecting low- and middle-income countries (LMIC) where healthcare access and disease control efforts are often constrained.[1] While TB control efforts have focused on high-risk populations, the growing burden of diabetes mellitus (DM) presents a compounding challenge.

Diabetes increases the risk of TB by approximately two-to threefold, and people with diabetes are more likely to have poor TB treatment outcomes, including delayed sputum conversion, increased relapse rates, and higher mortality.[2-4] Given the rising prevalence of diabetes, currently affecting an estimated 537 million adults worldwide, with the highest growth projected in LMIC, the convergence of these two epidemics poses a significant public health threat.[5] In the South African context, the intersection of TB and diabetes presents a significant public health concern. South Africa has one of the highest TB burdens globally, with an estimated incidence of 468 cases per 100,000 population in 2022 (WHO, 2024). The prevalence of diabetes is increasing, with the latest reported prevalence of 11% among South African adults in 2021.[5]

Recognizing this, the World Health Organization (WHO) recommends systematic TB screening for people with diabetes in high TB-burden settings as part of its collaborative framework for TB and DM control.[6] The recommended screening strategies typically include symptom-based screening algorithms and chest X-ray assessments.[7] However, the recommendations are based on data mostly derived from the general population and other risk groups such as contacts, and the performance of these screening tools among people with diabetes remains poorly understood.

To date, there have been limited studies on the accuracy of TB screening tools in people with diabetes. The clinical manifestations of TB in individuals with diabetes is known to differ from those without diabetes, with a higher probability of more severe presentations, cavitary lung lesions, and atypical chest X-ray findings.[8] In addition, co-morbidities commonly associated with diabetes and older age, such as non-TB lung pathology, may further compromise the accuracy of symptom-based or radiographic screening tools in this population. Therefore, it is critical to validate the accuracy of recommended screening tools in this population. A study in Pakistan evaluated the accuracy of computer-assisted chest X-ray reading (CAD4TB) for TB screening in people with diabetes.[9] The study reported a sensitivity ranging from 48.6% to 90.5%, depending on the threshold used, which was lower than the 91.0%–97.3% sensitivity observed in individuals with unknown diabetes status in the same setting. However, diabetic status in this study was determined by self-report and random blood glucose measurement. A recent review found additional four studies that assessed the accuracy of computer-assisted chest X-ray reading for TB screening. [10] However, in all of these studies, sputum samples were collected only from individuals with symptoms, which is likely to overestimate sensitivity. Further, the reported accuracy is not generalisable to systematic screening approaches where chest X-ray is performed regardless of symptoms. Moreover, no study has evaluated the accuracy of symptom-based TB screening in people with diabetes or how glycaemic control affects the performance of existing screening tools. Some studies have reported a higher prevalence of cavitary lesions in individuals with poorly controlled diabetes,[11, 12] which could potentially increase the sensitivity of chest X-ray; however, this has not been formally assessed.

To address this gap, we conducted a study to evaluate the sensitivity and specificity of TB screening tools, including TB symptoms and chest X-ray, among people with diabetes. We also evaluated how the accuracy of these tools differ by glycaemic control and other clinical factors. Our findings aim to inform evidence-based screening strategies for this high-risk population, ultimately contributing to improved TB detection and control efforts.

## Methods

### Study population and setting

This study utilized baseline data from a prospective cohort of adults with diabetes attending routine primary care in Khayelitsha, a subdistrict of Cape Town, South Africa. We consecutively enrolled adults aged 15 years and older diagnosed with any type of diabetes from the diabetes outpatient clinic within the Site B Community Health Centre.

Eligible individuals were those with a documented history of diabetes and current use of antidiabetic medications or those meeting diagnostic criteria (HbA1c ≥ 6.5% or fasting plasma glucose ≥7.0 mmol/L). Known HIV status was a requirement and both HIV-positive and HIV-negative individuals were included. The recruitment took place from 1 March 2022 to 31 November 2022.

We obtained written informed consent from all study participants. Ethical approval was obtained from the University of Cape Town Human Research Ethics Commitee (791/2021) and University College London (28119/001).

### Study investigations

We collected medical history and screened for TB symptoms, such as cough, fever, night sweats, and weight loss. All participants underwent chest X-ray. All participants, regardless of symptoms or chest X-ray findings, submitted a single sputum sample for testing by Xpert Ultra. Participants were also tested for HbA1c using venous blood samples.

### Definitions

We evaluated the performance of the following screening tools: any cough (cough of any duration); any TB symptoms, defined as at least one of any cough, fever, night sweats, or weight loss; and chest X-ray abnormality. Chest X-ray abnormalities were classified in two ways: TB-suggestive abnormalities defined as radiographic findings such as cavitation, consolidation, pleural effusion, or fibrosis that were indicative of TB and any lung abnormalities, which include other abnormalities, such as neoplastic lesions or chest wall abnormalities, not necessarily deemed suggestive of TB. Chest X-ray images were read independently by two trained physicians, with final interpretations based on consensus. Images were also evaluated using CAD4TB (Computer-Aided Detection for Tuberculosis; Delft Imaging Systems, ‘s-Hertogenbosch, the Netherlands), and the manufacturer-recommended threshold of ≥ 60 was used to define TB-suggestive abnormality. We also assessed combinations of any TB symptoms and chest X-ray abnormalities in parallel, using both classification schemes (i.e. TB suggestive abnormalities and any lung abnormalities). For these combination strategies, a positive screening result was defined as the presence of either TB symptoms or chest X-ray abnormalities, and a negative result as the absence of both.

A positive Xpert MTB/RIF Ultra test result served as the reference standard. Trace results were also considered positive in the primary analysis.

### Statistical analysis

First, we calculated the sensitivity, specificity, negative predictive value (NPV), and positive predictive value (PPV) for each screening criterion defined above. The corresponding 95% confidence intervals were estimated using the exact method.[13] We also assessed the accuracy of combining symptom-based and chest X-ray screening in parallel, Second, we assessed potential differences in specificity of the screening tools according to the following pre-defined factors: diabetic control (HbA1c <7% vs. HbA1c ≥7%), HIV status, previous history of TB, and sex. The specificity was estimated stratified by each factor, Logistic regression was used to assess whether specificity was associated with these factors.[14] We also examined the association between specificity by age, treating age as a continuous variable in the regression model. We intended to conduct similar analyses for sensitivity; however, due to the small number of individuals diagnosed with TB, we report sensitivity stratified by each factor where feasible, without regression analysis.

In sensitivity analyses, trace-positive Xpert MTB/RIF Ultra results were classified as negative. Multiple testing was not accounted for. All statistical analyses were conducted using STATA version 18.0 (StataCorp).

## Results

We enrolled 673 adults with diabetes. Of these, 11 were unable to produce sputum and one had an indeterminate Xpert Ultra result; therefore, 661 participants were included in the analysis (Figure 1). Among these, the median age was 53 years (interquartile range 47-60 years), with the majority being female (63.8%) (Table 1). Most participants (83.4%) had a poor glycaemic control defined as HbA1c ≥ 7%, and 17.4% were HIV positive. TB was diagnosed in nine (1.4%, 95% confidence interval [CI] 0.6-2.6%) of the participants and all of them were HIV negative. Chest X-ray results were missing for 84 participants (12.9%) without a TB diagnosis. Among 580 participants with chest X-ray results CAD4TB scores were missing for 49 participants (8.4%) without TB, including one participant with TB.

**Table 1.** Characteristics of participants.

|  | All (N= 661) | Non-TB (N=652) | TB (N=9) |
| --- | --- | --- | --- |
| Age (median [IQR]) | 53.00 [47.00, 60.00] | 53.00 [47.00, 60.00] | 63.00 [50.00, 68.00] |
| Gender = Male (%) | 239 (36.2) | 234 (35.9) | 5 (55.6) |
| Current smoker (%) | 25 (4.0) | 25 (4.1) | 0 (0.0) |
| Current alcohol use (%) | 124 (18.9) | 123 (19.0) | 1 (11.1) |
| BMI (%) |  |  |  |
| Underweight (BMI <18.5) | 2 (0.3) | 2 (0.3) | 0 (0.0) |
| Normal weight (BMI 18.5-24.9) | 94 (14.3) | 90 (13.9) | 4 (44.4) |
| Overweight (BMI 25.0-29.9) | 184 (28.0) | 181 (27.9) | 3 (33.3) |
| Obesity (BMI ≥30.0) | 377 (57.4) | 375 (57.9) | 2 (22.2) |
| Poor glycaemic control (HbA1c ≥ 7%) (%) | 504 (83.4) | 499 (83.7) | 5 (62.5) |
| HbA1c (median [IQR]) | 9.40 [7.70, 11.50] | 9.40 [7.70, 11.50] | 10.05 [6.75, 12.48] |
| HIV-positive (%) | 115 (17.4) | 115 (17.6) | 0 (0.0) |
| Previous history of TB (%) | 114 (17.2) | 111 (17.0) | 3 (33.3) |

IQR: interquartile range; BMI: body mass index; HIV: human immunodeficiency virus; TB:
tuberculosis

**Figure 1.**
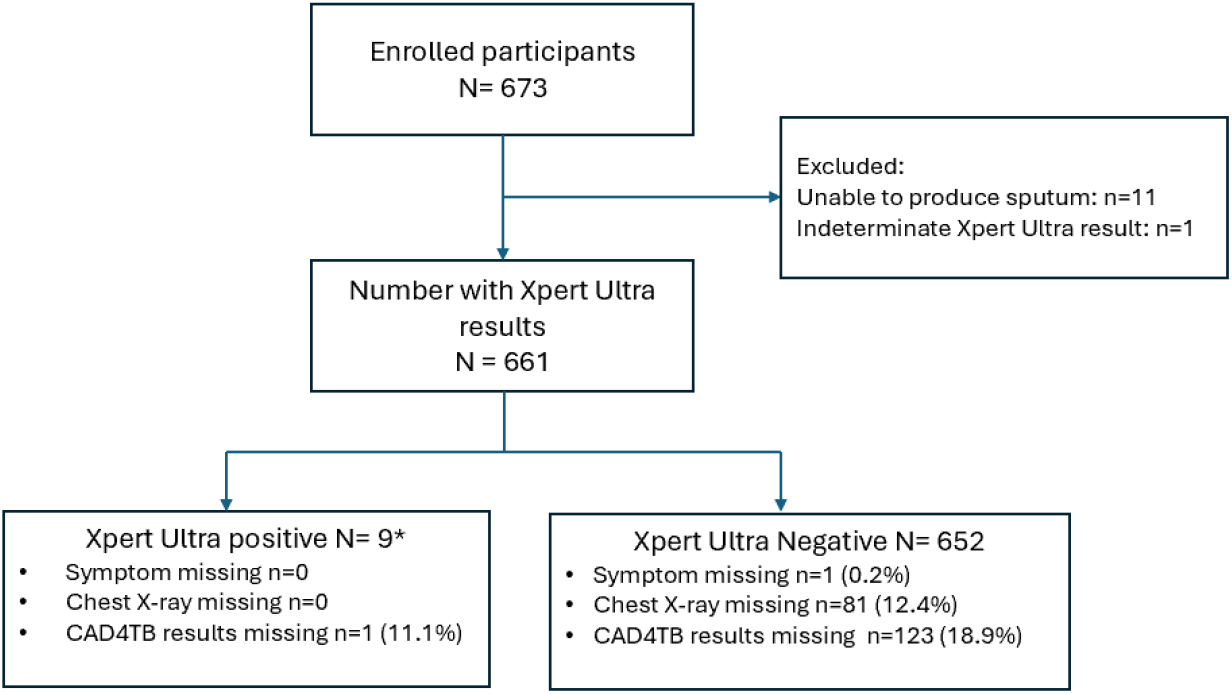
Flow diagram showing the numbers of patients studied. *Include two trace results, which were considered negative in sensitivity analysis. Alt text: Flow diagram of participant inclusion and test results. Of 673 enrolled participants, 12 were excluded, leaving 661 with Xpert Ultra results. Among these, 9 tested positive and 652 tested negative.

### Accuracy of TB symptoms and chest X-ray abnormality

Among nine people with TB, only two had any cough, resulting in a sensitivity of 22.2% (95% CI 2.8-60.0%) (Table 2). The specificity of any TB symptoms was high at 97.5% (95% CI 96.0-98.6%). The use of any TB symptoms did not increase the sensitivity, with a slight reduction of specificity (96.0%, 95% CI: 94.2–97.4%).

**Table 2.** Accuracy of symptom and chest X-ray screening.

| Screening tool | TP | FN | FP | TN | Sensitivity, % (95% CI) | Specificity, % (95% CI) | PPV, % (95% CI) | NPV, % (95% CI) |
| --- | --- | --- | --- | --- | --- | --- | --- | --- |
| Any cough | 2 | 7 | 16 | 635 | 22.2 (2.81-60.0) | 97.5 (96.0-98.6) | 11.1 (1.4-34.7) | 98.9 (97.8-99.6) |
| Any TB symptoms | 2 | 7 | 26 | 625 | 22.2 (2.81-60.0) | 96.0 (94.2-97.4) | 7.1 (0.9-23.5) | 98.9 (97.7-99.6) |
| CXR-TB suggestive abnormalities | 5 | 4 | 26 | 542 | 55.6 (21.2-86.3) | 95.4 (93.4-97.0) | 16.1 (5.5-33.7) | 99.3 (98.1-99.8) |
| CXR-Any lung abnormalities | 5 | 4 | 37 | 531 | 55.6 (21.2-86.3) | 93.5 (91.1-95.4) | 11.9 (4.0-25.6) | 99.3 (98.1-99.8) |
| CAD4TB score ≥ 60 | 3 | 5 | 12 | 517 | 37.5 (8.5-75.5) | 97.7 (96.1-98.8) | 20.0 (4.3-48.1) | 99.0 (97.8-99.7) |
| CXR-TB suggestive abnormalities and any TB symptoms | 5 | 4 | 51 | 517 | 55.6 (21.2-86.3) | 91 (88.4-93.2) | 8.9 (3.0-19.6) | 99.2 (98.0-99.8) |
| CXR- any lung abnormalities and any TB symptoms | 5 | 4 | 60 | 508 | 55.6 (21.2-86.3) | 89.4 (86.6-91.8) | 7.7 (2.5-17.0) | 99.2 (98.0-99.8) |

TP: true positive; FN: false negative; FP: false positive; TN: true negative; CXR: chest X-ray; CI: confidence interval; PPV: positive predictive value; NPV: negative predictive value; TB: tuberculosis

Chest X-ray had a higher sensitivity than TB symptoms at 55.6% (95% CI 21.2-86.3%) for both chest X-ray abnormalities suggestive of TB and any lung abnormalities. The specificity of chest X-ray abnormalities suggestive TB was similar to TB symptoms (95.4%, 95% CI 93.4-97.0%) while that of any lung abnormalities was slightly lower (93.5%, 95% CI 91.1-95.4%). CAD4TB had a slightly higher specificity (97.7%, 95% CI 96.1-98.8) and its sensitivity was 37.5% (95% CI 8.5-75.5). When using the combination of any positive TB symptoms or chest X-ray signs strategy, the sensitivity remained same whereas the specificity declined slightly (91%, 95% CI 88.4-93.2% for the combination of any TB symptoms and chest X-ray abnormalities suggestive of TB). The PPV was highest for chest X-ray abnormalities suggestive of TB.

In sensitivity analysis, where trace results were considered negative, the sensitivity of chest X-ray (for both TB-suggestive findings and any lung abnormalities) increased substantially to 71.4% (95% CI: 29.0–96.3%), while specificity remained largely unchanged (Table S1).

### Accuracy of TB symptoms and chest X-ray abnormalities stratified by clinical factors

The sensitivity of chest X-ray was higher among those with poor glycaemic control (HbA1c ≥7.0%;66.7%, 95% CI: 22.3–95.7%) compared to those with better control (HbA1c < 7.0%; 33.3%, 95% CI:0.8–90.6%), and higher in females (75.0%, 95% CI: 19.4–99.4%) than in males (40.0%, 95% CI: 5.3– 85.3%), although confidence intervals were wide and overlapping. Differences in sensitivity by HIV status could not be evaluated due to the absence of TB cases among HIV-positive individuals. The specificity of symptom-based screening remained consistent across clinical subgroups (Tables S2-S5).

Participants with a previous history of TB had a substantially lower specificity across all chest X-ray-based tools, including CAD4TB. For instance, specificity for chest X-ray abnormalities suggestive of TB was 87.6% (95% CI: 79.8–90.6%) among those with prior TB, compared to 97.2% (95% CI: 95.2–98.5%) in those without (p < 0.01). Similarly, the specificity of chest X-ray with any lung abnormalities was 84.8% (95% CI: 76.4–91.0%) vs. 95.5% (95% CI: 93.2–97.2%), respectively (p < 0.01).

A lower specificity was also observed among HIV-positive individuals for any lung chest X-ray detecting abnormalities (88.8%, 95% CI: 80.8–94.3%) compared to HIV-negative individuals (94.5%,95% CI: 92.0–96.4%; unadjusted p = 0.04), and in males compared to females (91.3%, 95% CI: 86.7–94.8% vs. 94.7%, 95% CI: 91.9–96.8%; unadjusted p = 0.012). However, when previous TB history was accounted for, there was no evidence specificity differed by HIV status (adjusted p=0.481) or sex (adjusted p=0.225), whereas prior TB history remained a significant predictor of reduced specificity. Likewise, older age was not significantly associated with a reduction of specificity (Table S6).

## Discussion

This study assessed the accuracy of different TB screening tools among people with diabetes in a high-burden setting, using a rigorous prospective design with systematic sputum collection regardless of symptoms or chest X-ray findings. Our findings contribute to optimising TB screening strategies for people with diabetes in high-burden settings.

The sensitivity of symptom-based screening alone was suboptimal, consistent with findings in other populations.[15] The estimated sensitivity of any TB symptoms was notably lower than that cited in the WHO systematic screening guidelines (22.2% vs. 71%).[7] Chest X-ray had a higher sensitivity than symptom screening but was also lower than reported in other populations. However, due to the small number of people with TB in our study, the sensitivity estimates were imprecise. Moreover, our study population consisted of people with diabetes already in routine care, likely screened for TB symptoms multiple times. As a result, people with symptomatic TB may have already been diagnosed prior to enrolment and screening in our study, leaving only individuals with minimal or no symptoms and those without notable chest X-ray abnormalities in our analysis, thereby reducing the sensitivity of the screening tools. On the other hand, the specificity of both symptom screening and chest X-ray was high, although specificity was slightly lower for chest X-ray based on human reading. Our study could not demonstrate increased sensitivity by combining symptom screening and chest X-ray, likely due to the small number of TB cases.

Our study found a significantly lower specificity of chest X-ray abnormalities in individuals with a history of TB compared to those without. This is likely due to persistent chest X-ray abnormalities from previous TB, leading to false positives. On the other hand, the specificity of the screening tools remained consistent across other clinical factors, such as glycemic control, which is reassuring.

However, the impact of these clinical factors on sensitivity remains unclear and requires further evaluation.

In this cohort of people with diabetes, the prevalence of TB was 1.4%, nearly double the prevalence in the general population (0.85%) as reported in a recent national survey.[16] This was observed despite male individuals, who are at higher risk for TB, accounting for less than half of our study population. These findings reinforce the importance of systematic TB screening in people with diabetes to prevent the morbidity and mortality associated with TB in this vulnerable group. Chest X-ray is likely the most efficient screening approach given its high PPV; however, reliance on Chest X-ray requires careful consideration of feasibility, accessibility, and cost, particularly in resource-limited settings.

The use of computer-assisted reading of chest X-rays may help address some of these challenges by reducing the need for specialist interpretation and enabling broader implementation in peripheral settings [17]. Furthermore, its specificity was higher than that of human reading, which could reduce the number of participants requiring confirmatory tests, although its impact on sensitivity could not be estimated with precision. The use of additional tools, such as C-reactive protein, may warrant exploration,[18] but data on its utility among people with diabetes are currently unavailable.

There are several limitations to our study. First, as previously noted, the small number of TB cases resulted in wide confidence intervals for sensitivity estimates and precluded an assessment of the impact of clinical factors on sensitivity. Nonetheless, our study allowed for a precise assessment of specificity, contributing new knowledge to the existing body of evidence. Second, our study was conducted in a single clinic, enrolling people with diabetes already in care. The findings may not be generalizable to other settings or to individuals with newly diagnosed diabetes, although the specificity estimates were similar across multiple clinical factors. Lastly, the reliance on sputum-based Xpert Ultra as the reference standard excludes extrapulmonary TB. Given that its sensitivity is not 100% and that only a single sputum sample was tested, some people with TB may have been missed. Additionally, Xpert Ultra may have detected dead bacilli in individuals without symptoms or X-ray abnormalities, potentially underestimating the sensitivity of the screening tools, as suggested by the sensitivity analysis: although with wide confidence intervals.

## Conclusion

Our study underscores the limitations of symptom-based TB screening in people with diabetes and highlights the potential of chest X-ray to improve sensitivity, although it remained suboptimal.

Additionally, the specificity of chest X-ray declined in people with previous TB. The high prevalence of TB observed in our study among people with diabetes calls for further exploration of additional screening tools to enhance TB case detection while ensuring feasibility and cost-effectiveness. The findings of our study need to be validated in other settings, partly due to the imprecise sensitivity estimates. Such studies will contribute to refining screening strategies to address the growing burden of diabetes in high TB-burden settings and support efforts to achieve the goal of ending TB.

## Acknowledgment

Y.H. and N.J. conceived and designed the study. A.T. led participant recruitment. L.B. and W.C. served as study clinicians and performed chest X-ray interpretation. R.G. was the study coordinator, and A.J. managed the study data. S.G., N.O.-D., L.L.S., and L.T. contributed to study implementation.

T.D. provided statistical advice. Y.H. and N.J. conducted the statistical analysis. N.J. and Y.H. drafted the manuscript. M.X.R. and R.J.W. provided overall supervision. All authors critically reviewed the manuscript, contributed important intellectual content, and approved the final version.

## Data availability

Requests for anonymized individual-level data underlying this study will be reviewed by CIDRI-Africa to determine whether the request is subject to confidentiality and data protection obligations. Data that can be shared will be made available upon reasonable request and subject to a data-sharing agreement.

## References

1. World Health Organization. Global TB Report, 2024. Geneva, Switzerland: WHO, 2024. https://www.who.int/teams/global-tuberculosis-programme/tb-reports/global-tuberculosis-report-2024 Accessed 31 October 2024).

2. Al-Rifai RH, Pearson F, Critchley JA, Abu-Raddad LJ. Association between diabetes mellitus and active tuberculosis: A systematic review and meta-analysis. PLoS One 2017; 12(11): e0187967.

3. Hayashi S, Chandramohan D. Risk of active tuberculosis among people with diabetes mellitus: systematic review and meta-analysis. Trop Med Int Health 2018; 23(10): 1058–70.

4. Restrepo BI. Diabetes and Tuberculosis. Microbiol Spectr 2016; 4(6).

5. IDF Diabetes Atlas 2021. https://diabetesatlas.org/data/en/.

6. World Health Organization. WHO operational handbook on tuberculosis. Module 6: tuberculosis and comorbidities, third edition. https://iris.who.int/bitstream/handle/10665/380063/9789240103276-eng.pdf?sequence=1 (accessed 9 April 2025).

7. World Health Organization. WHO consolidated guidelines on tuberculosis. Module 2: screening – systematic screening for tuberculosis disease. https://apps.who.int/iris/bitstream/handle/10665/340255/9789240022676-eng.pdf (accessed 9April 2025).

8. van Crevel R, Critchley JA. The Interaction of Diabetes and Tuberculosis: Translating Research to Policy and Practice. Trop Med Infect Dis 2021; 6(1).

9. Habib SS, Rafiq S, Zaidi SMA, et al. Evaluation of computer aided detection of tuberculosis on chest radiography among people with diabetes in Karachi Pakistan. Sci Rep 2020; 10(1): 6276.

10. Emoru RD, Mrema LE, Ntinginya NE, Biraro IA, van Crevel R, Critchley JA. Accuracy of computer-aided chest x-ray interpretation for tuberculosis screening in people with diabetes mellitus: A systematic review. Trop Med Int Health 2025; 30(5): 323–31.

11. Chiang C-Y, Lee J-J, Chien S-T, et al. Glycemic Control and Radiographic Manifestations of Tuberculosis in Diabetic Patients. PLoS One 2014; 9(4): e93397.

12. Meng F, Lan L, Wu G, et al. Impact of diabetes itself and glycemic control status on tuberculosis. Front Endocrinol (Lausanne) 2023; 14: 1250001.

13. Newcombe RG. Two-sided confidence intervals for the single proportion: comparison of seven methods. Stat Med 1998; 17(8): 857–72.

14. Bell C, Goss M, Birstler J, et al. Assessment of potential factors associated with the sensitivity and specificity of Sofia Influenza A+B Fluorescent Immunoassay in an ambulatory care setting. PLoS One 2022; 17(5): e0268279.

15. van’t Hoog A, Viney K, Biermann O, Yang B, Leeflang MMG, Langendam MW. Symptom- and chest-radiography screening for active pulmonary tuberculosis in HIV-negative adults and adults with unknown HIV status. Cochrane Database of Systematic Reviews 2022; (3).

16. Moyo S, Ismail F, Van der Walt M, et al. Prevalence of bacteriologically confirmed pulmonary tuberculosis in South Africa, 2017-19: a multistage, cluster-based, cross-sectional survey. Lancet Infect Dis 2022; 22(8): 1172–80.

17. Qin ZZ, Ahmed S, Sarker MS, et al. Tuberculosis detection from chest x-rays for triaging in a high tuberculosis-burden setting: an evaluation of five artificial intelligence algorithms. The Lancet Digital Health 2021; 3(9): e543–e54.

18. Ruperez M, Shanaube K, Mureithi L, et al. Use of point-of-care C-reactive protein testing for screening of tuberculosis in the community in high-burden settings: a prospective, cross-sectional study in Zambia and South Africa. The Lancet Global Health 2023; 11(5): e704–e14.

